# EIRsurvival: Deep Learning-based time-to-event analysis on high-dimensional genotype and multi-omics data

**DOI:** 10.1101/2025.03.05.25323428

**Authors:** Justus F. Gräf, Arnor I. Sigurdsson, Chiara Rohrer, Torben Hansen, Simon Rasmussen

## Abstract

**Motivation:** Time-to-event data in disease occurrence is often right-censored, requiring survival models for accurate predictions. While deep learning advancements have extended traditional Cox models, current approaches do not allow modeling on individual-level, large-scale genotype data. Scalable models integrating genetic and clinical data could enhance precision medicine and disease prediction.

**Results:** We introduce EIRsurvival, a deep learning-based tool designed to perform time-to-event analyses on millions of SNPs from large, individual-level cohorts. We applied EIRsurvival to predict time-to-diagnosis for eight diseases in the UK Biobank, utilizing 1.13M genetic variants from 487,027 individuals. The model achieved C-indices between 0.55 and 0.68 using genetic data alone, with performance improving to 0.67–0.96 when integrating multi-omics data. Automated feature attribution analysis confirmed biologically relevant feature associations.

**Availability and Implementation:** EIRsurvival is available through the eir-dl framework at github.com/arnor-sigurdsson/EIR, with documentation accessible at eir.readthedocs.io/en/latest/other/01_survival_genotypes.html. The tool is optimized for GPU acceleration, facilitating efficient training on high-dimensional data.

**Contact:** Simon Rasmussen

**Supplementary Information:** Supplementary information is available online.

## Introduction

Data on disease occurrences in individuals oftentimes comes in a right-censored format due to the nature of how diagnosis data is collected. To handle this type of data, survival models have been developed. A widely used example is the Cox Proportional Hazards model, which represents an individual’s hazard as a scaled version of a baseline hazard function, based on the individual’s feature values (Cox 1972). As a commonly used metric, the C-index indicates the survival model’s ability to rank individuals by their risk (Harrell, Lee, and Mark 1996). While a C-index of 0.5 indicates random ranking of the individuals, a C-index of 1 means that the model was able to rank the individuals in perfect order.

Availability of large scale omics data in individual level cohorts opened new applications for such models to identify associations between omics features and a time-to-diagnosis (time-to-event). For example, recent studies in the UK Biobank showed that proteomics data can predict the time-to-diagnosis for various diseases using a Cox-Proportional Hazard model (Carrasco-Zanini et al. 2024). Additionally, using a similar approach, Deng et al., identified 168,100 associations between proteins and disease occurrence (Deng et al. 2025).

However, these approaches are usually only applied to a limited number of features as input (i.e., 1-20 different proteins in a single model), while disease prediction could benefit from integrating many clinical and omics features in the same model. Additionally, the low dimensionality makes it challenging to capture complex effects between the features that potentially impact time-to-diagnosis. Deep Learning (DL) implementations have been developed to increase complexity in time-to-event models, which are extensively reviewed by Wiegrebe et al. (Wiegrebe et al. 2024). Importantly, while some of these models can work with omics data with thousands of features, very high dimensional data like individual-level genotypes currently cannot be included in time-to-event analyses. Yet, integrating genetics with clinical data becomes increasingly important and has potential to advance precision medicine (Ashley 2016). Additionally, the growing sizes of population-based genetic cohorts require efficient models that scale to large sample sizes (Bycroft et al. 2018).

Previously, we developed EIR, a Deep Learning framework that includes the Genome-local Net (GLN) model for genomic predictions (Arnór I. Sigurdsson et al. 2023). Based on this, we here introduce EIRsurvival, a DL-based tool to perform time-to-event analyses using millions of SNPs from hundreds of thousands of patients. It allows for integration of genetics with other (tabular) omics modalities or clinical features and provides extensive downstream analyses and model explainability. EIRsurvival builds the foundation for large-scale analyses of time-to-event data and can be used to identify novel associations, for example between genetic variants and the time-to-diagnosis of diseases.

EIRsurvival is available through the eir-dl framework at github.com/arnor-sigurdsson/EIR. A documentation of how to use EIRsurvival to predict time-to-event from genetics data can be found under eir.readthedocs.io/en/latest/other/01_survival_genotypes.html.

## Results

EIRsurvival implements Cox Proportional Hazards models with Efron approximation for handling tied survival times (Efron 1977). The framework accepts pre-processed genotypes from plink_pipelines (Sigurdsson et al., 2023), which is a custom pipeline to convert standard genotype data into NumPy arrays (Harris et al. 2020). EIRsurvival automatically adjusts to the size of the input genotype data, and can predict time-to-event for any individual-level outcome, such as disease diagnosis.

To demonstrate its use, we predicted the time-to-diagnosis for 8 diseases (AFF: Atrial Fibrillation and Flutter, AMI: Acute Myocardial Infarction, ASH: Asthma, GT: Gout, HT: Hypertension, HTD: Hyperthyroidism, T1D: Type 1 diabetes, T2D: Type 2 diabetes) in the UK Biobank from 1.13M genetic variants in 487,027 individuals across ancestries. The individuals were randomly split into training (n=438,326), validation (n=24,345) and test sets (n=24,356) before training. EIRsurvival trained on genotype data as input reached C-indices between 0.55 (HTD) and 0.68 (T2D) on the independent test set (n=24,356) (**Figure 1b**). To gain insights into the learned attributes, EIRsurvival includes an automated calculation of the model attribution for each variant using an integrated gradients method (Sundararajan, Taly, and Yan 2017). For example, variants with the highest attribution in the time-to-diagnosis model for AFF were located in the PITX2 loci which has known association to AFF (Nielsen et al. 2018). Similarly, for T2D, variants with the highest attributions were located in the TCF7L2 locus, which has well-known associations to T2D (Del Bosque-Plata et al. 2021). These analyses showed that EIRsurvival uses disease-relevant genetic variants to make time-to-event predictions. Time-to-event models are often represented as survival curves that show the survival probability over time. EIRsurvival produces a survival curve for each model that contains high-risk, low-risk, median risk predictions and the Kaplan-Meier Estimate (**Supplementary Figure 1a**) (Rich et al. 2010).

**Figure 1:**
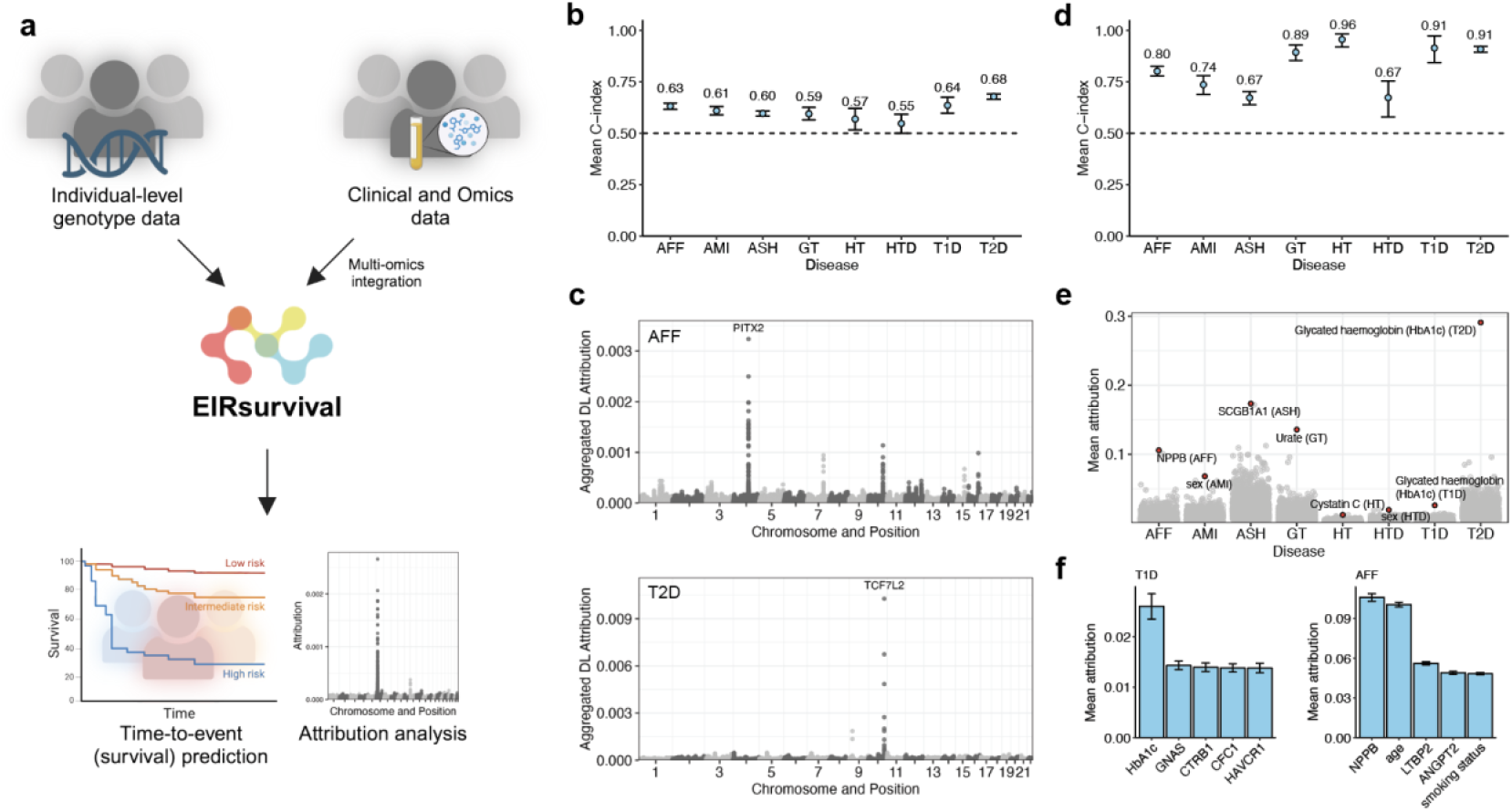
**a)** Summary of EIRsurvival functionality. **b)** Mean C-indices of 1000 bootstraps for time-to-event prediction (from time of birth) of 8 diseases (AFF: Atrial Fibrillation and Flutter, AMI: Acute Myocardial Infarction, ASH: Asthma, GT: Gout, HT: Hypertension, HTD: Hyperthyroidism, T1D: Type 1 diabetes, T2D: Type 2 diabetes) in the UK Biobank from genotype data of 1.13M variants. The models were trained in a 5-fold cross validation on variants from 462,671 individuals (train=438,326, validation=24,345) and tested on an independent test set of 24,356 individuals. The error bars represent the 95% Confidence Interval (CI) of the bootstrapped (n=1000) C-indices. **c)** Mean aggregated DL attribution across 5-folds on the test set for each of the 1.13M genetic variants for AFF and T2D. Known AFF and T2D loci with high DL attributions were labeled. **d)** Mean C-indices of 1,000 bootstraps for time-to-event prediction (from assessment center visit) of 8 diseases from 1.13M variants, 30 biomarkers, 2,012 plasma protein measurements, age, sex, BMI, smoking status and number of medications in use. The error bars represent the 95% CIs of the bootstrapped C-indices. **e)** Mean bootstrapped (n=1,000) DL attributions across 5-folds on the test set for each disease and input feature (excluding genetic variants). The input feature with the highest attribution was labeled for each disease. **f)** Top 5 DL attributions for T1D and AFF.

An important feature of EIRsurvival is that it can also integrate other, high-dimensional omics data. To demonstrate this, we integrated 1.13M SNPs, 30 biomarkers, 2,012 plasma protein measurements (Olink) as well as age, sex, BMI and smoking status from up to 52,632 individuals (dependent on disease). These models were trained on ∼42,000 individuals, and validated and tested on ∼5,200 individuals, depending on the disease (**Supplementary Information**). When predicting the time-to-diagnosis for the same eight diseases, the multi-omics EIRsurvival models reached performances between 0.67 (HTD) and 0.96 (HT) (**Figure 1d**). This corresponded to performance improvements between 0.07 (ASH) and 0.38 (HT) compared to the model trained on genotypes only. Here, the inbuilt attribution estimation offers explainability of the model that makes it possible to identify associations between clinical/omics features and time-to-diagnosis. For example, we found the highest feature attribution for T1D and T2D for the biomarker HbA1c, which is a biomarker for long-term blood glucose levels and used to diagnose and monitor diabetes (**Figure 1e**). Additionally, CTRB1 plasma levels were among the top attributions for T1D, which has been shown to be causally associated with T1D risk (Yazdanpanah et al. 2022). Furthermore, known risk factors for AFF, such as age and smoking status, were among the highest attributions for AFF (**Figure 1f**) (Elliott et al. 2023). This demonstrated that EIRsurvival was capable of identifying relevant clinical and omics associations with time-to-diagnosis of diseases from thousands of features. Additionally, the integration of genotype with clinical and omics features highly improved the predictive power of the time-to-event models.

EIRsurvival is optimized for running on GPUs, which can significantly reduce computing time, especially on high-dimensional data. For example, we trained a single fold for each disease on genetics data on either CPUs or GPUs and measured the computing time for 10,000 mini-batch iterations of the underlying GLN model (**Supplementary Figure 1b**). We found that models trained on GPUs were between 14-fold and 23-fold faster than trained on CPUs.

## Discussion

EIRsurvival provides a novel DL-based implementation of time-to-event models that efficiently scales to very high dimensional data. In particular, it can model individual level genotype data, which complements existing methods that have so far been limited to tabular omics data (Wiegrebe et al. 2024). We demonstrated its use by predicting the time-to-diagnosis of 8 diseases in 487,027 individuals of the UK Biobank. Expectedly, additional integration of multi-omics and clinical features significantly improved predictions, while demonstrating the multi-modal abilities of EIRsurvival.

DL has proven to effectively capture complex effects in high-dimensional data that influences traits and complex diseases (Elgart et al. 2022; Arnór I. Sigurdsson et al. 2023; Arnor I. Sigurdsson et al. 2024). Due to its implementation through the GLN of the EIR framework, EIRsurvival can likely capture those complex effects where present and thereby improve time-to-event prediction of complex traits. The integrated downstream analyses that EIR provides improve the explainability of the model and can help identify new associations between features and time-to-event. Notably, we showed that EIRsurvival identifies known disease-associated variants as well as clinical feature associations from millions of variants and thousands of clinical/omics features.

Furthermore, DL models greatly benefit from the use of GPUs, which offer substantial speed up over traditional CPU-based computations due to their highly parallel architecture. While EIRsurvival can be run on CPUs, it is optimized to run on GPUs, which makes it more feasible to train on high-dimensional data.

In summary, EIRsurvival addresses a gap in survival analysis by enabling the direct incorporation of genotype data, offering new opportunities to uncover single genetic variant contributions to time-to-event predictions. We anticipate it will serve as a valuable tool for researchers to perform risk prediction and discover new associations of omics traits with health and disease.

## Acknowledgements

The present analyses were conducted under UK Biobank data application number 32683. The UK Biobank received ethical approval from the North West - Haydock Research Ethics Committee (21/NW/0157).

## Competing Interest Statement

S.R. is the founder and owner of BioAI and has performed consulting for Sidera Bio ApS. The remaining authors declare no competing interests.

## Funding

S.R., J.F.G., and T.H. were supported by the Novo Nordisk Foundation (NNF23SA0084103).

J.F.G. was supported by a research grant from the Danish Cardiovascular Academy, which is funded by the Novo Nordisk Foundation, grant number NNF20SA0067242 and The Danish Heart Foundation.

## Data availability

UK Biobank genotype, proteomics and covariate data is available to approved researchers through the UK Biobank (https://www.ukbiobank.ac.uk/enable-your-research/apply-for-access).

## Supplementary Information

### Documentation and instructions

EIRsurvival is an implementation of CPH models in the already existing EIR framework (Arnór I. Sigurdsson et al. 2023). A tutorial of how to use the EIRsurvival implementation to predict time-to-events from genotypes can be found under eir.readthedocs.io/en/latest/other/01_survival_genotypes.html. A more comprehensive documentation and more tutorials on the EIR framework are available under https://eir.readthedocs.io/en/latest/index.html.

### Time-to-diagnosis analysis of 8 diseases in the UK Biobank

ICD10 codes from the hospital in-patient records in the UK Biobank were mapped to phecodes using the PheWAS catalog (Denny et al. 2013). The time-to-diagnosis for each individual and disease was calculated in years from their year of the birth. Individuals that died or did not get diagnosed with the disease by 2020-12-31 were censored. For the multi-omics integration, the time-to-diagnosis was calculated in days from the date of assessment center visit and individuals that got diagnosed before or in the 180 days following the assessment center visit were removed from the analysis. This led to slight differences in training, validation and test set numbers between the diseases for the multi-omics models.

### Genotype and proteomics data processing

For the demonstration of EIRsurvival on genetics data, filtered HapMap3 genotypes were reused from a previous project (Gräf et al., in preparation). In short, 1.3M HapMap3 variants were filtered based on –geno 0.05 and –hwe 1e-45 calculated on a subset of the UKB-Pharma proteomics cohort (n=44,883), resulting in 1.13M variants included in the analysis. For the Olink plasma proteomics measurements that were used as input in the multi-omics models only protein measurements were included that had at least one protein-phenotype association in the proteome-phenome atlas (n=2,012) (Deng et al. 2025). All features that were integrated together with the genotypes are listed in **Supplementary Table 1** together with their UKB field IDs.

### C-index and DL attributions

The C-indices were bootstrapped for each fold with n=1,000 bootstraps from the test set predictions and the mean and 95% confidence interval was calculated across all bootstraps across the five folds. The DL attribution was averaged across the five folds for each variant. For the clinical and omics models, the mean attributions and 95% CIs were calculated across 1000 bootstraps and five folds.

**Supplementary Figure 1:**
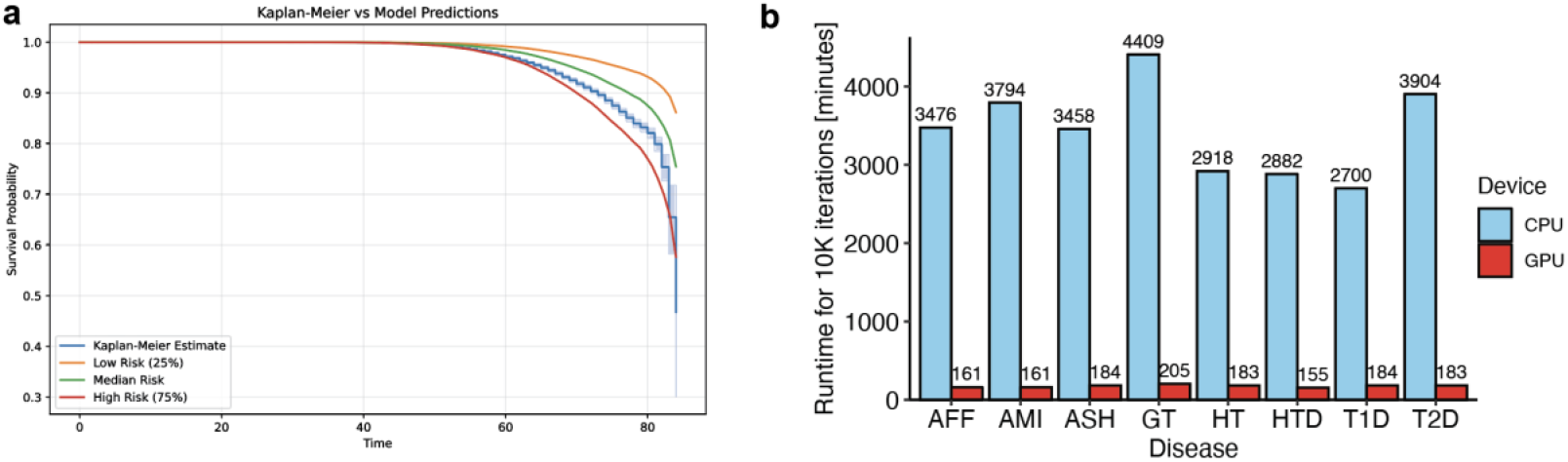
**a)** Example of a survival plot produced by EIRsurvival for each model representing the probability of survival (that an event does not occur) over time (in years). Here a survival plot for T2D is shown. **b)** Runtime in minutes for 10,000 training iterations of the Genome-Local Net model run on CPU (5 core, AMD EPYC 7543) or GPU (NVIDIA A100) for a single fold with a batch size of 64 (10,000 iterations corresponding to 640,000 samples).

## References

Ashley, Euan A. 2016. “Towards Precision Medicine.” Nature Reviews. Genetics 17 (9): 507–22.

Bycroft, Clare, Colin Freeman, Desislava Petkova, Gavin Band, Lloyd T. Elliott, Kevin Sharp, Allan Motyer, et al. 2018. “The UK Biobank Resource with Deep Phenotyping and Genomic Data.” Nature 562 (7726): 203–9.

Carrasco-Zanini, Julia, Maik Pietzner, Jonathan Davitte, Praveen Surendran, Damien C. Croteau-Chonka, Chloe Robins, Ana Torralbo, et al. 2024. “Proteomic Signatures Improve Risk Prediction for Common and Rare Diseases.” Nature Medicine 30 (9): 2489–98.

Cox, D. R. 1972. “Regression Models and Life-Tables.” Journal of the Royal Statistical Society.

Series B, Statistical Methodology 34 (2): 187–202.

Del Bosque-Plata, Laura, Eduardo Martínez-Martínez, Miguel Ángel Espinoza-Camacho, and Claudia Gragnoli. 2021. “The Role of in Type 2 Diabetes.” Diabetes 70 (6): 1220–28.

Deng, Yue-Ting, Jia You, Yu He, Yi Zhang, Hai-Yun Li, Xin-Rui Wu, Ji-Yun Cheng, et al. 2025. “Atlas of the Plasma Proteome in Health and Disease in 53,026 Adults.” Cell 188 (1): 253–71.e7.

Denny, Joshua C., Lisa Bastarache, Marylyn D. Ritchie, Robert J. Carroll, Raquel Zink, Jonathan D. Mosley, Julie R. Field, et al. 2013. “Systematic Comparison of Phenome-Wide Association Study of Electronic Medical Record Data and Genome-Wide Association Study Data.” Nature Biotechnology 31 (12): 1102–10.

Efron, Bradley. 1977. “The Efficiency of Cox’s Likelihood Function for Censored Data.” Journal of the American Statistical Association 72 (359): 557.

Elgart, Michael, Genevieve Lyons, Santiago Romero-Brufau, Nuzulul Kurniansyah, Jennifer A. Brody, Xiuqing Guo, Henry J. Lin, et al. 2022. “Non-Linear Machine Learning Models Incorporating SNPs and PRS Improve Polygenic Prediction in Diverse Human Populations.” Communications Biology 5 (1): 856.

Elliott, Adrian D., Melissa E. Middeldorp, Isabelle C. Van Gelder, Christine M. Albert, and Prashanthan Sanders. 2023. “Epidemiology and Modifiable Risk Factors for Atrial Fibrillation.” Nature Reviews. Cardiology 20 (6): 404–17.

Gräf, Justus F., Arnór I. Sigurdsson, Jonas Meisner, Chiara Rohrer, Benjamin M. Neale, Torben Hansen, and Simon Rasmussen. n.d. “Prediction of Plasma Proteins in the UK Biobank.” In Preparation.

Harrell, F. E., Jr, K. L. Lee, and D. B. Mark. 1996. “Multivariable Prognostic Models: Issues in Developing Models, Evaluating Assumptions and Adequacy, and Measuring and Reducing Errors.” Statistics in Medicine 15 (4): 361–87.

Harris, Charles R., K. Jarrod Millman, Stéfan J. van der Walt, Ralf Gommers, Pauli Virtanen, David Cournapeau, Eric Wieser, et al. 2020. “Array Programming with NumPy.” Nature 585 (7825): 357–62.

Nielsen, Jonas B., Rosa B. Thorolfsdottir, Lars G. Fritsche, Wei Zhou, Morten W. Skov, Sarah E. Graham, Todd J. Herron, et al. 2018. “Biobank-Driven Genomic Discovery Yields New Insight into Atrial Fibrillation Biology.” Nature Genetics 50 (9): 1234–39.

Rich, Jason T., J. Gail Neely, Randal C. Paniello, Courtney C. J. Voelker, Brian Nussenbaum, and Eric W. Wang. 2010. “A Practical Guide to Understanding Kaplan-Meier Curves.” Otolaryngology--Head and Neck Surgery : Official Journal of American Academy of Otolaryngology-Head and Neck Surgery 143 (3): 331–36.

Sigurdsson, Arnor I., Justus F. Gräf, Zhiyu Yang, Kirstine Ravn, Jonas Meisner, Roman Thielemann, Henry Webel, et al. 2024. “Non-Linear Genetic Regulation of the Blood Plasma Proteome.” medRxiv. 10.1101/2024.07.04.24309942.

Sigurdsson, Arnór I., Ioannis Louloudis, Karina Banasik, David Westergaard, Ole Winther, Ole Lund, Sisse Rye Ostrowski, et al. 2023. “Deep Integrative Models for Large-Scale Human Genomics.” Nucleic Acids Research 51 (12): e67.

Sundararajan, Mukund, Ankur Taly, and Qiqi Yan. 2017. “Axiomatic Attribution for Deep Networks.” http://arxiv.org/abs/1703.01365.

Wiegrebe, Simon, Philipp Kopper, Raphael Sonabend, Bernd Bischl, and Andreas Bender. 2024. “Deep Learning for Survival Analysis: A Review.” Artificial Intelligence Review 57 (3). 10.1007/s10462-023-10681-3.

Yazdanpanah, Nahid, Mojgan Yazdanpanah, Ye Wang, Vincenzo Forgetta, Michael Pollak, Constantin Polychronakos, J. Brent Richards, and Despoina Manousaki. 2022. “Clinically Relevant Circulating Protein Biomarkers for Type 1 Diabetes: Evidence From a Two-Sample Mendelian Randomization Study.” Diabetes Care 45 (1): 169–77.

